# Designing AI-powered healthcare assistants to effectively reach vulnerable populations with health care services: A discrete choice experiment among South African university students

**DOI:** 10.1101/2025.01.30.25321409

**Authors:** A Zheng, L Long, C Govathson, C Chetty-Makkan, S Morris, D Rech, MP Fox, S Pascoe

## Abstract

**Introduction:** South African young adults are at increased risk for HIV acquisition and other non-communicable diseases and face significant barriers to accessing healthcare services. The rapid development of artificial intelligence (AI), in particular AI-powered healthcare assistants (AIPHA), presents a unique opportunity to increase access to health information and linkage to healthcare services and providers. While successful implementation and uptake of such tools require understanding user preferences, limited understanding of these preferences exist. We sought to understand what preferences are important to university students in South Africa when engaging with a hypothetical AIPHA to access health information using a discrete choice experiment.

**Methods:** We conducted an unlabeled, forced choice discrete choice experiment among adult South African university students through Prolific Academic, an online research platform, in 2024. Each choice option described a hypothetical AIPHA using eight attribute characteristics (cost, confidentiality, security, healthcare topics, language, persona, access, services). Participants were presented with ten choice sets each comprised of two choice options and asked to choose between the two. A conditional logit model was used to evaluate preferences.

**Results:** 300 participants were recruited and enrolled. Most participants were Black, born in South Africa, heterosexual, working for a wage, and a mean age of 26.5 years (SD: 6.0). Results from the discrete choice experiment identified that language, security, and receiving personally tailored advice were the most important attributes for AIPHA. Participants strongly preferred the ability to communicate with the AIPHA in any South African language of their choosing instead of only English and to receive information about health topics specific to their context including information on clinics geographically near them. Results were consistent when stratified by sex and socioeconomic status.

**Conclusions:** Participants had strong preferences for security and language which is in line with previous studies where successful uptake and implementation of such health interventions clearly addressed these concerns. These results build the evidence base for how we might engage young adults in healthcare through technology effectively.

## Introduction

Accessing appropriate sexual and reproductive health (SRH) services for adolescents and young adults is critical to prevent unintended pregnancies, the transmission of sexually transmitted infections (STIs), sexually transmitted diseases (STDs), and HIV. A key component of being able to access SRH services is knowledge and education on SRH. In South Africa many young adults, including university students, have low SRH knowledge which contributes to high-risk sexual behavior (not using a condom, multiple partners, and early sexual debut), and in turn increases their risk of unintended pregnancies, transmission of STIs, STDs, and HIV.(1–4) This is particularly concerning as HIV is highly prevalent and one of the leading causes of morbidity and mortality in South Africa and achieving epidemic control hinges on ensuring HIV prevention services reach individuals who would benefit from it most. (5–8) This population’s limited SRH knowledge may help explain, in part, why they are underrepresented in HIV prevention services despite being disproportionately affected by HIV.(1,4,9–11) University students represent a subset of the young adult population who may be at an increased risk due to recently having moved away from home to attend university and exposure to behaviors that might increase their vulnerability to STIs, STDS, and HIV. (8,9,12,13) However, attendance at a university also presents a unique opportunity to provide a greater degree of health services, including SRH care and services, as university students are physically concentrated on campus.(8,12–14) Despite such an opportunity, health services in particular SRH services including HIV care and prevention, on campus are often limited particularly at under resourced institutions.(8,14) Furthermore, South African university students have a high prevalence of mental health conditions such as anxiety, depression alcohol and substance use, which can in turn impact their willingness and ability to access in-person health services.(15–17) Being able to reach young adult South Africans, provide them with appropriate health information, identify their healthcare needs and rapidly steer them towards appropriate health services remains a critical gap in improving their overall health and reaching HIV epidemic control.

Youth and young adults experience substantial barriers in identifying and using health care services.(14,15,18–20) Common barriers that have been identified include low health literacy, long wait times at health facilities, inability to take time away from classes to attend health facilities, and the attitudes and behaviors of healthcare workers.(14,15,21) Long wait times may be a stronger deterrent for young adults who are not ill and are only seeking information and preventative services. Similarly, healthcare worker attitude may be particularly important for young adults who are seeking SRH services that are often stigmatized. Recent advances in artificial intelligence (AI), in particular large-language models leveraged into AI-powered healthcare assistants (AIPHA), present an opportunity to address some of these barriers and help countries achieve the United Nations Sustainable Development Goals, in particular Goal 3.7, which is focused on ensuring universal access to SRH services as well as information and education on SRH.(22–25) Within the health sector this has involved AI tools that can perform a broad range of tasks from providing health information to risk screening, and scheduling appointments to providing advanced diagnostic advice and capabilities.(23,24,26–31) Preliminary work suggests that AIPHAs could be particularly suited to educating clients, gathering relevant medical history, and referring to appropriate services especially if the topics are taboo or potentially stigmatizing, like sexual history and HIV status. However, a critical component of their success will be presenting and designing the AIPHAs so they meet the specific needs and preferences of the end users.

Despite the exponential growth of AI-powered health care applications, there is a dearth of data to identify characteristics that end users of these health applications value and would drive uptake.(32,33). Cost has been found to be a characteristic that affects South African healthcare users decisions in the public sector and as most South Africans have to pay for internet data this may be important for AI-powered applications.(34,35) Similarly, if we draw parallels from the concerns clients have raised with in person health interactions, any characteristics of an application related to confidentiality (i.e. password protection, storage of data), the interaction experience (i.e. friendliness and personality of the tool persona), and the breadth of knowledge (i.e. conditions covered, knowledge of relevant services) may be important for decision making.(32,33,36) Structural questions, such as how the health application is accessed (i.e. online, downloaded application) and in what language, are likely to influence use in places like South Africa where most of the population only has access to basic smart phone capabilities and there is great language diversity (11 official spoken South African languages plus languages from neighboring countries).(34) While there is a growing body of literature to support the importance of these characteristics in these AI-driven tools, to our knowledge there has been no attempt to evaluate the importance of these characteristics alongside one another in decision making around the use of AI-powered healthcare assistants (AIPHA).(23,24,29,31,37–39)

Given that the use of AIPHAs is becoming widespread across many settings and applications and has great potential in addressing gaps in the HIV care cascade in healthcare settings where resources are limited, we attempted to identify which attributes (or characteristics) users value in deciding whether to use such an assistant. As context and who are the end users are key factors that can influence preferences and uptake, we enrolled potential end users (young adults) within the local environment of interest (attending university).(39) Specifically, we evaluated the stated preferences of South African tertiary students for various attributes of a hypothetical AIPHA to support their healthcare decision making and access, including sexual and reproductive health. This was done using a discrete choice experiment (DCE) which allowed us to determine the student’s preference for a specific attribute when deciding across various AIPHAs.(40)

## Methods

### Advisory Committee

An eight-person advisory committee was created to provide technical oversight and local context when considering the selection of attributes (the characteristics of a choice option) to include in the DCE. This committee included the research implementation team; South African researchers with experience in discrete choice experiments, HIV prevention and working with populations at increased vulnerability; and AI technology experts with implementation experience in South Africa and the region. The advisory committee was involved in all major decisions regarding study design, analysis and interpretation and provided guidance on attributes and their associated levels (the values that the characteristic can assume).(40,41) For example, “language” might be an attribute and the levels it could take on might be “English only,” “English and slang,” and “All local languages.”

### Development of Attributes and Levels and Ranking Survey

Based on the literature, we created a provisional list of modifiable attributes (characteristics) of a hypothetical AIPHA likely to be important for South African university students.(8,13–15,20,31,38,39,42–44) The advisory committee reviewed and augmented the list to create 13 overarching attributes (Supplemental Table 1) that they believed might be important when developing and utilizing an AIPHA. DCE best practice suggests approximately eight attributes with at most four levels per attribute to ensure that participants can cognitively weigh each choice and not be overburdened by the different choice options. Utilizing the Prolific Academic, an online research platform, we then conducted a survey asking 30 adult university students (≥ 18 years old) in South Africa to rank these 13 attributes of an AI tool from most important to least important when engaging with an AIPHA. Use of Prolific Academic in behavioral research such as DCEs has been well-established.(45,46) To create a single “super”-list that was reflective of all participants, we conducted a rank aggregation using the Cross-Entropy Monte Carlo algorithm.(47,48) See Supplemental Text 1 for more details on this algorithm. Based on this ranking and input from the advisory committee we identified the eight attributes likely to be the most important. The final eight attributes and the possible levels were presented to the South African research team to ensure appropriateness and clarity in language (Table 1). These finalized attributes and levels were used to design the DCE survey instrument.

**Table 1.**
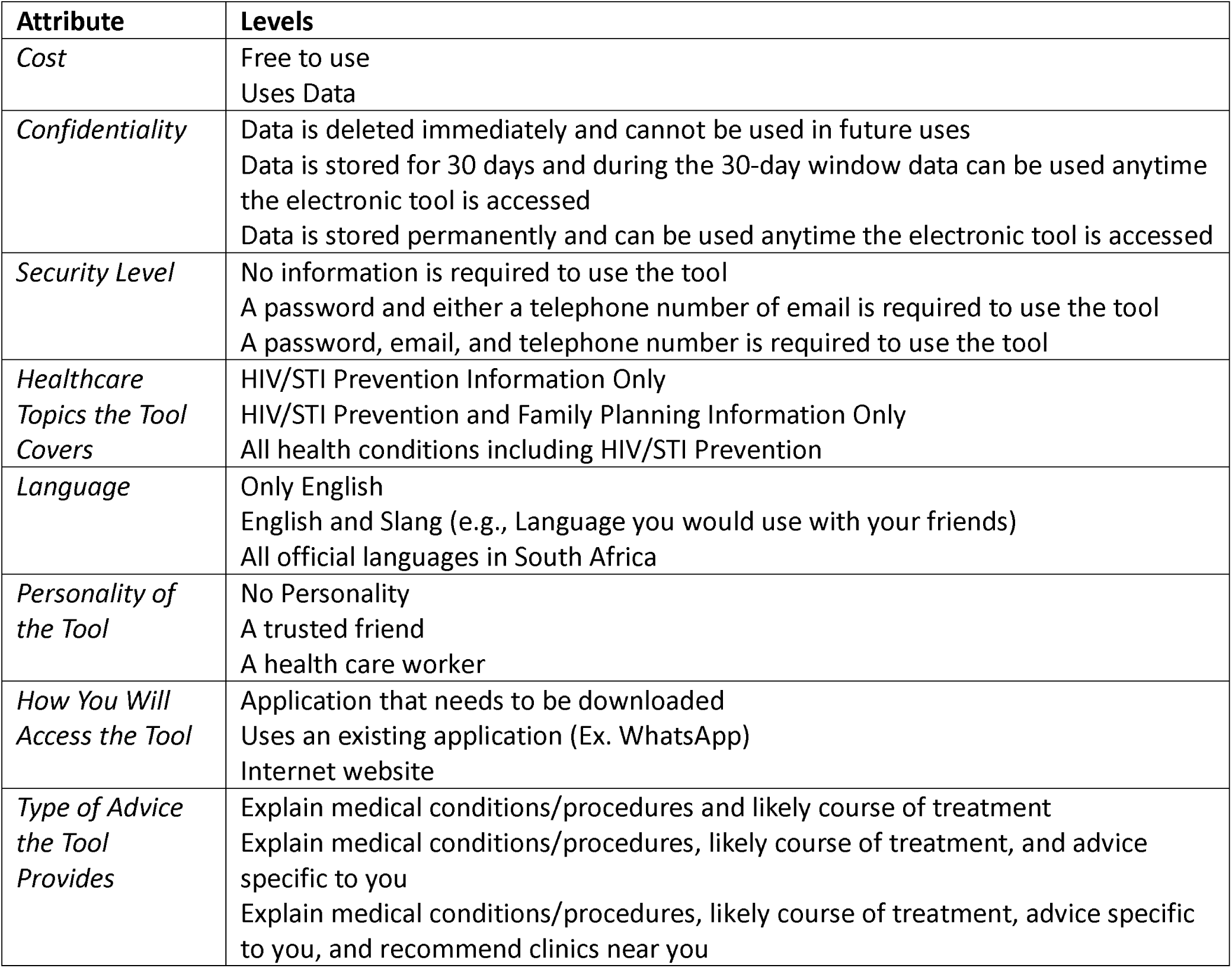
Attributes and Levels of the Discrete Choice Experiment

**Table 2.** Baseline demographics stratified by sex and overall (N = 300)

|  | **Male**<br>(n = 150) | **Female**<br>(n = 150) | **Total**<br>(N = 300) |
| --- | --- | --- | --- |
| **Age (Years)** | 26.5 (5.3) | 26.6 (6.7) | 26.5 (6.0) |
| **Ethnicity** |  |  |  |
| *Black* | 130 (86.7) | 127 (84.7) | 257 (85.7) |
| *Mixed* | 6 (4.0) | 11 (7.3) | 17 (5.7) |
| *White* | 8 (5.3) | 6 (4.0) | 14 (4.7) |
| *Asian* | 1 (0.7) | 2 (1.3) | 3 (1.0) |
| *Other* | 4 (2.7) | 3 (2.0) | 7 (2.3) |
| *Decline to Answer* | 1 (0.7) | 1 (0.7) | 2 (0.7) |
| **Nationality** |  |  |  |
| *South African* | 137 (91.3) | 141 (94.0) | 278 (92.7) |
| *Other* | 13 (8.7) | 9 (6.0) | 22 (7.3) |
| **Country of Birth** |  |  |  |
| *South Africa* | 137 (91.3) | 140 (93.3) | 277 (92.3) |
| *Other* | 13 (8.7) | 10 (6.7) | 23 (7.7) |
| **Province** |  |  |  |
| *Eastern Cape* | 9 (6.0) | 12 (8.0) | 21 (7.0) |
| *Free State* | 5 (3.3) | 8 (5.3) | 13 (4.3) |
| *Gauteng* | 83 (55.3) | 82 (54.7) | 165 (55.0) |
| *KwaZulu-Natal* | 16 (10.7) | 14 (9.3) | 30 (10.0) |
| *Limpopo* | 9 (6.0) | 5 (3.3) | 14 (4.7) |
| *Mpumalanga* | 3 (2.0) | 5 (3.3) | 8 (2.7) |
| *North West* | 8 (5.3) | 9 (6.0) | 17 (5.7) |
| *Northern Cape* | 2 (1.3) | 0 | 2 (0.7) |
| *Western Cape* | 15 (10.0) | 15 (10.0) | 30 (10.0) |
| **Institution Type** |  |  |  |
| *University* | 128 (85.3) | 127 (84.7) | 255 (85.0) |
| *T-VET College* | 8 (5.3) | 8 (5.3) | 16 (5.3) |
| *Other Tertiary Institution* | 14 (9.3) | 15 (10.0) | 29 (9.7) |
| **Year at Tertiary Institution** |  |  |  |
| *First* | 25 (16.7) | 22 (14.7) | 47 (15.7) |
| *Second* | 33 (22.0) | 25 (16.7) | 58 (19.3) |
| *Third* | 37 (24.7) | 47 (31.3) | 84 (28.0) |
| *Fourth* | 36 (24.0) | 39 (26.0) | 75 (25.0) |
| *Greater than Fourth* | 19 (12.7) | 17 (11.3) | 36 (12.0) |
| **Gender** |  |  |  |
| *Female* | 0 | 149 (99.3) | 149 (49.7) |
| *Male* | 149 (99.3) | 0 | 149 (49.7) |
| *Transgender* | 1 (0.7) | 1 (0.7) | 2 (0.6) |
| **Sexual Orientation** |  |  |  |
| *Heterosexual* | 144 (96.0) | 126 (84.0) | 270 (90.0) |
| *Bisexual* | 2 (1.3) | 17 (11.3) | 19 (6.3) |
| *Gay* | 3 (2.0) | 0 | 3 (1.0) |
| *Lesbian* | 0 | 5 (3.3) | 5 (1.7) |
| *Other* | 1 (0.7) | 2 (1.4) | 3 (1.0) |
| **Socioeconomic Status<sup>1</sup>** |  |  |  |
| *Low* | 54 (36.0) | 44 (29.3) | 98 (32.7) |
| *Medium* | 46 (30.7) | 45 (30.0) | 91 (30.3) |
| *High* | 50 (33.3) | 61 (40.7) | 111 (37.0) |
| **Currently Working for a Wage** | 89 (59.3) | 107 (71.3) | 196 (65.3) |
| **Own a Car** | 61 (40.7) | 62 (41.3) | 123 (41.0) |
| **Employ Domestic Workers** | 45 (30.0) | 58 (38.7) | 103 (34.3) |
| **Medical Aid Coverage** |  |  |  |
| *Yes* | 55 (36.7) | 69 (46.0) | 124 (41.3) |
| *No* | 90 (60.0) | 77 (51.3) | 167 (55.7) |
| *Decline to Answer* | 1 (0.7) | 1 (0.7) | 2 (0.7) |
| *I do not know* | 4 (2.7) | 3 (2.0) | 7 (2.3) |
| **Place Last Sought Medical Care** |  |  |  |
| *Private Setting<sup>2</sup>* | 74 (49.3) | 90 (60.0) | 164 (54.7) |
| *Public Setting<sup>3</sup>* | 72 (48.0) | 52 (34.7) | 124 (41.3) |
| *Other<sup>4</sup>* | 3 (2.0) | 4 (2.7) | 7 (2.3) |
| *Decline to Answer* | 0 | 1 (0.7) | 1 (0.3) |
| *I do not know* | 1 (0.7) | 3 (2.0) | 4 (1.3) |

<sup>1</sup>Based on the following questions: owning a car, employing a domestic work, and having medical aid coverage. Low: Answer no to all 3 questions; Medium: Answer yes to only 1 of the 3 questions; High: Answer yes to at least 2 of the 3 questions
<sup>2</sup>Private doctor, hospital, clinic, nurse, or chemist
<sup>3</sup>Public health clinic or hospital
<sup>4</sup>Includes traditional healer

### Survey Instrument Design and Implementation

The survey instrument consisted of a set of sociodemographic questions as well as the DCE choice sets (10 choice sets per participant). The sociodemographic questions were adopted from standardized questions used in South Africa and included questions on age, sex, sexual orientation, province of residence, socioeconomic status, and medical insurance status. We conducted an unlabeled, forced choice DCE where participants were asked to choose between two alternatives (A and B), where each alternative was described using the eight attributes set at different levels. Figure 1 shows one potential choice set with two alternatives.

**Figure 1.**
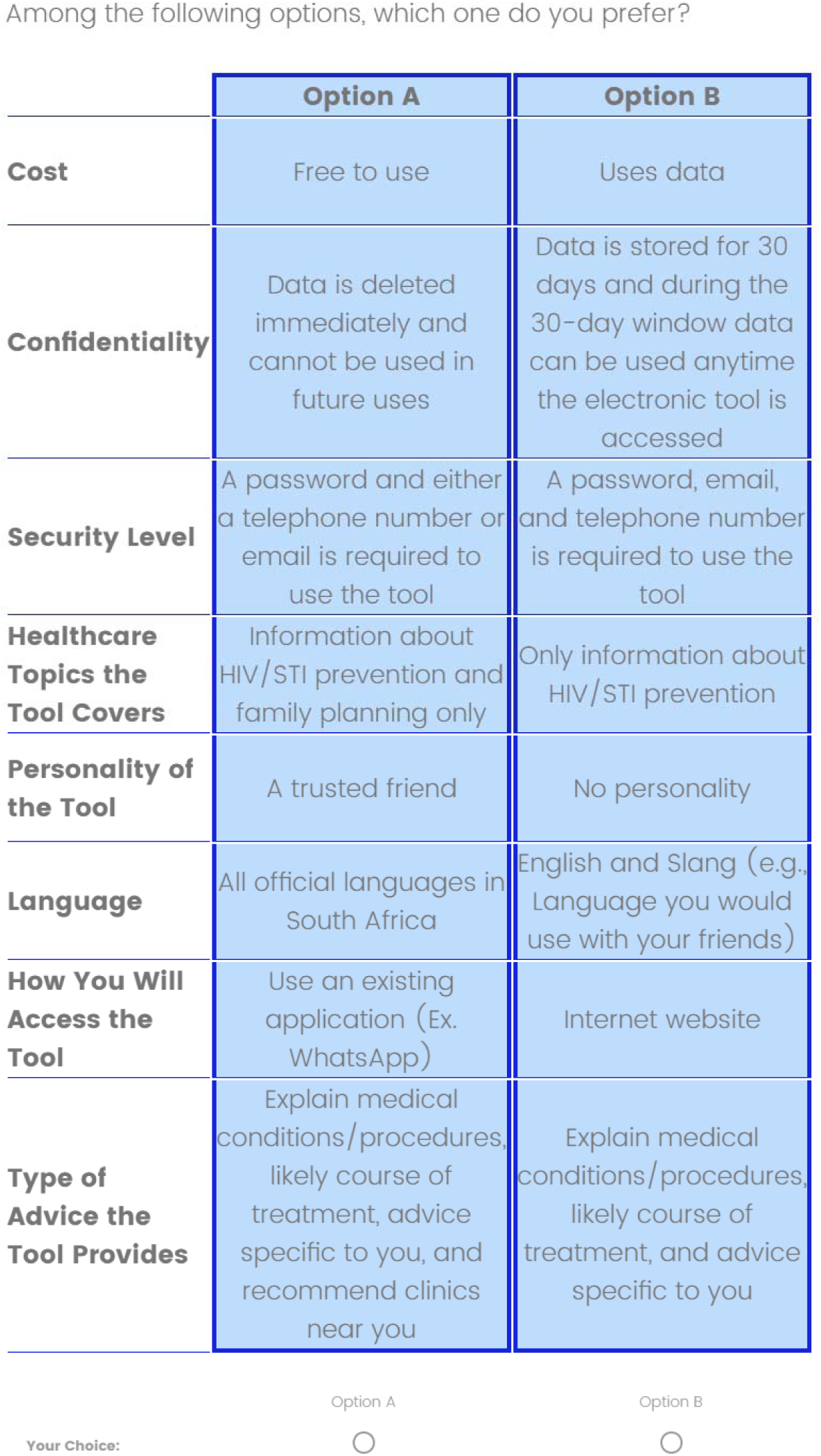
Example choice set that is presented to the participants.

We used Stata to generate a *D-efficient* experimental design to maximize the information contained in each choice set using the modified Fedorov algorithm.(49) Supplemental Tables 2 and 3 demonstrate that the choice sets were balanced and orthogonal. The experimental design had 18 choice sets from which we created two blocks. To prevent respondent fatigue and cognitive overload, each participant saw only nine unique choice sets.(40,50) We included a duplicate choice set to measure internal consistency, each participant therefore completed 10 choice sets. Participants were randomized 1:1 to each block. The survey was administered in English and implemented using Qualtrics, an online survey platform.(51) Prior to completing the DCE participants watched a brief video that explained the attributes, levels and data collection activities for the DCE.

### Sample Size Estimation

To determine the appropriate sample size the formula from Johnson and Orme (Equation 1) was used as an approximate in the absence of prior preference estimates.(52) Based on this formula, with ten choice sets and four levels, a minimum sample size of 100 was required. This was then increased to 150 based on literature that suggests substantial increases in the precision of estimates up to a sample size of 150.(50,53) Given that there may be differences in preferences by sex, we doubled the sample size for a final sample size of 300 to allow for stratification by sex assigned at birth.

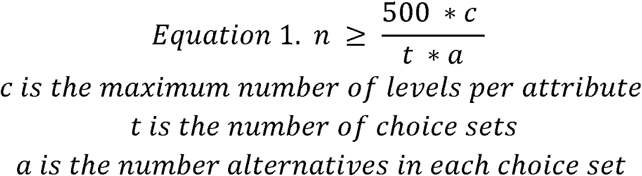

### Screening and Recruitment

Participants for the ranking survey and the main DCE were recruited through Prolific Academic. To be eligible for the ranking survey and the DCE, participants had to be adults (≥18 years old), reside in South Africa, be enrolled at a tertiary-level institution, be fluent in English, and have a Prolific Academic approval rating of ≥ 80% (e.g., a score that indicates a participant’s reliability and quality as a participant in a study). Only participants who met the inclusion/exclusion criteria were offered an opportunity to complete the survey on Prolific Academic. A description of the study was provided to eligible participants which included the expected completion time, the number of participants being recruited, how it could be completed (i.e., tablet, computer, phone, etc.), and reimbursement. Individuals then could choose to complete or not complete the survey. Participants who elected to participate were redirected to Qualtrics to complete informed consent and the study survey. Participants who completed the initial ranking survey to determine which attributes to include in the DCE were not eligible to participate in the main DCE. The survey sample was balanced by sex assigned at birth as recorded on the Prolific Academic database.

### Piloting of Survey Instrument

We piloted the study among ten participants to ensure the DCE could be completed within 30 minutes and that there were no issues with study procedures such as the instructional video not playing nor with the wording of the DCE. As there were no issues identified and therefore no changes made to the instrument, the pilot sample was included in the analytic sample.

### Ethics, Informed Consent, and Reimbursement

The study protocol was reviewed and approved by the Human Research Ethics Committee of the University of Witwatersrand (231115) and the Boston University Medical Center Institutional Review Board (H-44483). Both approved a waiver of written informed consent so that participants could indicate consent online through an informed consent document shared with them on Qualtrics. The use of Prolific Academic meant that all participants were anonymous to the research team and no identifying information was provided. Participants were reimbursed for their participation at a rate of $12.00/hour (∼South African ZAR200/hour) as recommended by Prolific Academic.(54)

### Statistical Analysis

We used descriptive statistics to describe sociodemographic characteristics. Given that this was a forced choice, unlabeled DCE, a conditional logit model with dummy coding to analyze the choice data was conducted.(55–57) Analyses were stratified by sex and socioeconomic status. A rudimentary measure of socioeconomic status levels based on available data were based on three standard questions related to vehicle ownership, employment of household help, and medical insurance coverage. Participants who responded “No” to all three questions were defined as low, to two questions as medium, and to one or none as high socioeconomic status. We conducted the following four separate sensitivity analyses: 1) excluded those who completed the DCE in < 10 minutes as the survey was intended to take 20-30 minutes to complete, 2) excluded those who were the fastest 10% of all participants, 3) excluded those who did not pass the internal consistency check (e.g., the choice they chose for the duplicate choice set did not match the original choice set), and 4) using the repeat choice set instead. Results from the sensitivity analysis were compared to the primary analysis which included the full sample with the repeat choice set excluded.

## Results

### DCE Survey Participant Characteristics

Table 1 presents baseline demographic characteristics of the 300 participants. The mean age was 26.5 years (SD: 6.0), and most participants were Black, born in South Africa, enrolled at a university, heterosexual, and currently working for a wage. The survey took a median of 18.0 minutes (Interquartile Range [IQR]: 14.7-21.3) to complete. Internal consistency was acceptable with 215 respondents out of 300 choosing the same choice in the duplicate choice set as they did in the original choice set (71.8%).

### Primary and Stratified Analyses

Participants showed a strong preference for an AIPHA that securely stored their information by allowing them to create a password protected account with personal identifiers included (Odds Ratio (OR): 1.71, 95% Confidence Interval (CI): 1.50, 1.94; Figure 2; Supplemental Table 4) compared to an AIPHA that included no account information or personal identification. They also had a strong preference for tools that allowed the use of any of the official South African languages (OR: 1.80 95% CI: 1.60, 2.02), or at least English in combination with local slang and vernacular (OR: 1.37, 95% CI:1.22, 1.54) compared to only being in English. Giving the AIPHA a personality of either a healthcare worker (OR: 1.48, 95% CI:1.30, 1.68) or trusted friend (OR: 1.35, 95% CI:1.18, 1.54) was also preferred to having a chatbot agent with no personality. AIPHAs that could be used to discuss all health conditions (OR: 1.40, 95% CI: 1.24, 1.57) or at least a combination of HIV/STI prevention information and family planning (OR: 1.21, 95% CI: 1.06, 1.38) were preferred compared to an AIPHA only focused on HIV/STI prevention. AIPHAs that offered tailored advice and able to direct the client to clinics near them were preferred to those tools that only explained the medical conditions more generically. It was also preferred if these tools could be accessed through existing platforms like WhatsApp (OR: 1.49, 95% CI: 1.32, 1.68) or a website (OR: 1.14, 95% CI: 1.01, 1.29) rather than through a bespoke application. Having a cost associated with using the AIPHA (OR: 0.81, 95% CI: 0.74, 0.88) or an AIPHA that stored their questions and answers permanently (OR: 0.80, 95% CI: 0.70, 0.90) were potential deterrents to using an AIPHA.

**Figure 2.**
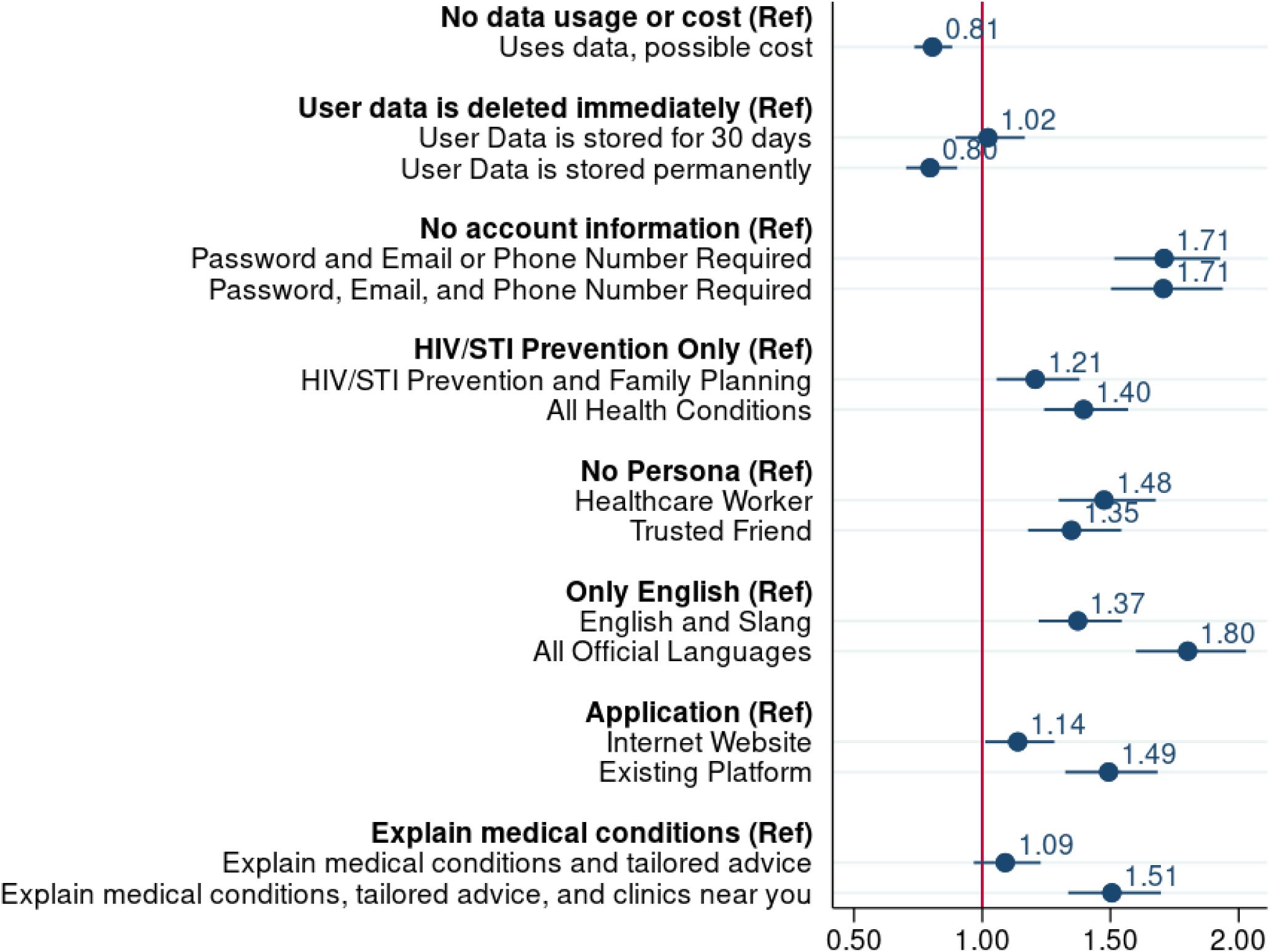
Primary findings from the Discrete Choice Experiment using a conditional logit model with dummy coding (N = 300)

### Sensitivity Analyses

Sensitivity analyses 1-3 demonstrated that findings were consistent (did not charge the order or relative size) with the primary analyses (Supplemental Figures 3-5). This suggests that speed with which the survey was completed or the fact that a participant had discordant results on the repeated choice set did not alter the overall results of the study. When using the repeat choice set instead (sensitivity analysis 4), the only notable change was participants had no preference in using a website over a bespoke application (OR: 1.00, 95% CI: 0.89, 1.13; Supplemental Figure 6).

## Discussion

Our study evaluated and measured preferences among South Africa university students for certain characteristics in an AIPHA. There is currently limited literature on the characteristics AIPHA should include given that only recently advancements in large-language models (LLMs) have allowed for the development of these assistants. Based on results from this study, having an AIPHA that used local languages, was password protected and had secure account information, and was able to provide tailored, personalized health advice including information on where to access nearby clinics, were the characteristics most strongly preferred among South African university students. These characteristics align with preferences for accessing health care services more broadly (i.e. in traditional in person health care settings) in South Africa as reflected in the growing body of literature exploring stated preferences for health service access. (58–60)

The strong preference to “converse” with an AIPHA in a local language aside from English, even when participants were proficient in English, highlights not only the importance of language diversity but the challenge faced by health care providers that serve very diverse populations. Supporting language diversity, whether in written health care material, with a healthcare provider or when communicating with an AIPHA, speaks directly to comprehension of health information and equitable access to health care services.(61) This is especially salient for tools that are powered by LLMs which to date have used English and so communication in African languages tends to be limited or more prone to mistakes.(62) Furthermore, AIPHA may need the ability to recognize and respond to conversations that utilize multiple languages as respondents may not only use their local language but can switch between languages within a single conversation. Work is being done by companies such as Meta to create AI models using African languages and slang.(63) A related concern is that much of the data utilized to train LLMs come from high-income countries and has been found to perpetuate biases especially against people from marginalized communities, people of lower socioeconomic status, non-Western identities such as those from the African continent, and people who experience greater difficulties in accessing care.(62,64–66) Therefore, any use of LLMs within the South African healthcare context must take this into account as it has been well-established that advancements in technology for health has the potential to advance access to health but may also exacerbate health disparities and inequities.(67–70)

The overlapping concerns of data security on phones (i.e. passwords, user accounts) and confidentiality of data (i.e. retention and sharing of data) when shared with an online platform were characteristics that might influence South African tertiary students’ decision to use an AIPHA to seek healthcare information and services. This is particularly important in the South African context where the prevalence of HIV is high and discussion of health topics with an AIPHA may carry the potential risk of unintended disclosure and result in stigma and discrimination. These findings are consistent with other studies that have found potential users of AI, including those using AI-powered assistants, express concerns regarding the security and privacy of the information they capture using those tools.(28,29,31,38) However, that research has also found that users may be more likely to use an AI conversational agent when it comes to accessing information on sensitive and stigmatized topics potentially because they perceive AI to be safer and more confidential than a healthcare provider and also more welcoming and less judgemental of their decisions and behaviors.(26,27,38) This suggests that if users concerns around security and data privacy can be met, an AIPHA may facilitate these critical conversations leading to appropriate and effective provision of health information.

As expected, any characteristics which increased the burden on the user for accessing the AIPHA would likely be a deterrent to using the tool (i.e. requiring a specific or new application to be downloaded or incurring a cost such as the need to acquire data). This is similar to existing literature that has identified cost and accessibility as barriers to access in-person healthcare services and predictive of successful uptake of healthcare interventions as a whole.(29,31,39) In South Africa, mobile devices are often shared between friends and within families so using existing applications with security features to ensure privacy, like WhatsApp which has the ability to lock chats, may help to ensure an individual’s information and discussions are kept private and confidential and prevent others from accessing their health-related chats.(71) Finally the participants expressed a preference for information and advice that was tailored to them (i.e. specific to their own medical condition, context and location). Evidence suggests that personalized medicine increases patient engagement and leads to better disease prevention.(72) Furthermore, participants expressed a stronger preference in being able to access information on health conditions beyond HIV/STI prevention information suggesting there is a desire for an AIPHA to serve as a “one-stop-shop” where participants can access information on multiple health related topics. This supports evidence that patients desire an integrated service delivery approach when it comes to seeking and receiving health care. Implementation of integrated service delivery is often challenging when funding for innovation, like AIPHA, is tied to specific service.

Our analysis has several limitations. First our study may not be representative of all young adult student populations as we recruited our study population from Prolific Academic, an online research platform. While the platform has been demonstrated to be appropriate for enrolling participants in behavioral research, users of the platform self-select into it and may be of higher socio-economic status with more exposure to technology and possibly information than the general population. Second, while our results when stratified by SES remained consistent with our primary findings, these results may not be adequately powered as we did not intentionally enroll equal numbers across each strata meaning that there were less than 100 participants in some of our categories (e.g., low and medium SES). Finally, our composite measure of SES was based on standardized household questions and may not accurately reflect and capture all facets of SES in this South African context. Specifically, they measured attributes that would generally be associated with a moderate to high level of socio-economic status in this setting and could be indicative of where the student stayed (i.e. with parents or student residence).

## Conclusions

In the rapidly changing landscape of AI powered innovation in healthcare, our work highlights the need to understand which characteristics of technology delivery are important for our end users – without this the full potential of this technology might never be realized. The findings show South African tertiary students accessing health information place a high priority on being able to communicate freely in their own language and the ability to protect themselves by managing access and data flow with an AIPHA. This needs to be balanced against their desire to receive personalized advice about health conditions and information on nearby clinics which requires access to their data and location. This work builds on the evidence base for engaging young adults in healthcare through technology effectively, but further work needs to be done to better understand whether these preferences vary for young adults outside of the tertiary education setting and if these preferences translate into uptake and ultimately improved health outcomes.

## Contributors Statement

Study conceptualization: AZ, LL, MPF, SP

Data curation, analysis and visualization: AZ, LL, CG

Software: AZ, LL, CG

Data analysis: AZ, LL, CG

Study supervision: LL, MPF, SP

Writing of initial draft: AZ

Review and editing: All coauthors

Access and verification of all data reported in the study: AZ, LL

All authors are responsible for the decision to submit for publication.

## Data Sharing

Data is not publicly available but may be provided in a de-identified format upon reasonable request.

## Acknowledgements

We would like to thank the participants for their time and contributions.

## Supplemental Tables and Figures Table of Contents

**Supplemental Table 1.** The 13 characteristics and examples of each characteristic identified by the advisory team

| **Characteristic** | **Example** |
| --- | --- |
| *Confidentiality* | What happens to the data that you enter (Stored Forever, Deleted Immediately etc.)? |
| *Modality* | How do you access the tool (Application, Through existing application like WhatsApp, Website etc.)? |
| *Persona of the Tool* | Does the tool have a personality? (Health Care Worker, Family Member, etc.) |
| *Availability* | Can the tool be accessed at any time? (24/7, Clinic Hours) |
| *Ability to Communicate with a Real Person* | Can you talk to a real healthcare worker through the tool? (No, Get a call back, Direct Connection, etc.) |
| *Language* | Can you only speak English with the tool? (Yes, English but can understand local slang, Bilingual) |
| *Cost* | Does the tool require data to use or is it data free? |
| *Privacy/Security Level* | How secure is the tool (Nothing, Password, Requires email, etc.)? |
| *Provide Real Time Clinical Recommendations* | Does the tool offer specific medical advice to you or can it only give general information (Only gathers clinical information, Can explain medical conditions, Offers personal insights) |
| *Audio/Voice to Text* | Is there an option to interact with tool that does not require typing? (No, Audio and Voice to Text) |
| *Location Services* | Does the tool give you information about clinics specific to your location? (No, you can ask based on a specific location, Yes you have the option of it using your location)? |
| *Relationship* | Is the tool able to build a relationship over multiple sessions? (No, Yes - Can remember and remind about appointments or adherence) |
| *Services Provided* | What health topics can the tool talk about? (HIV/STI Only, HIV/STI/Family Planning, All Health) |

**Supplemental Table 2.**
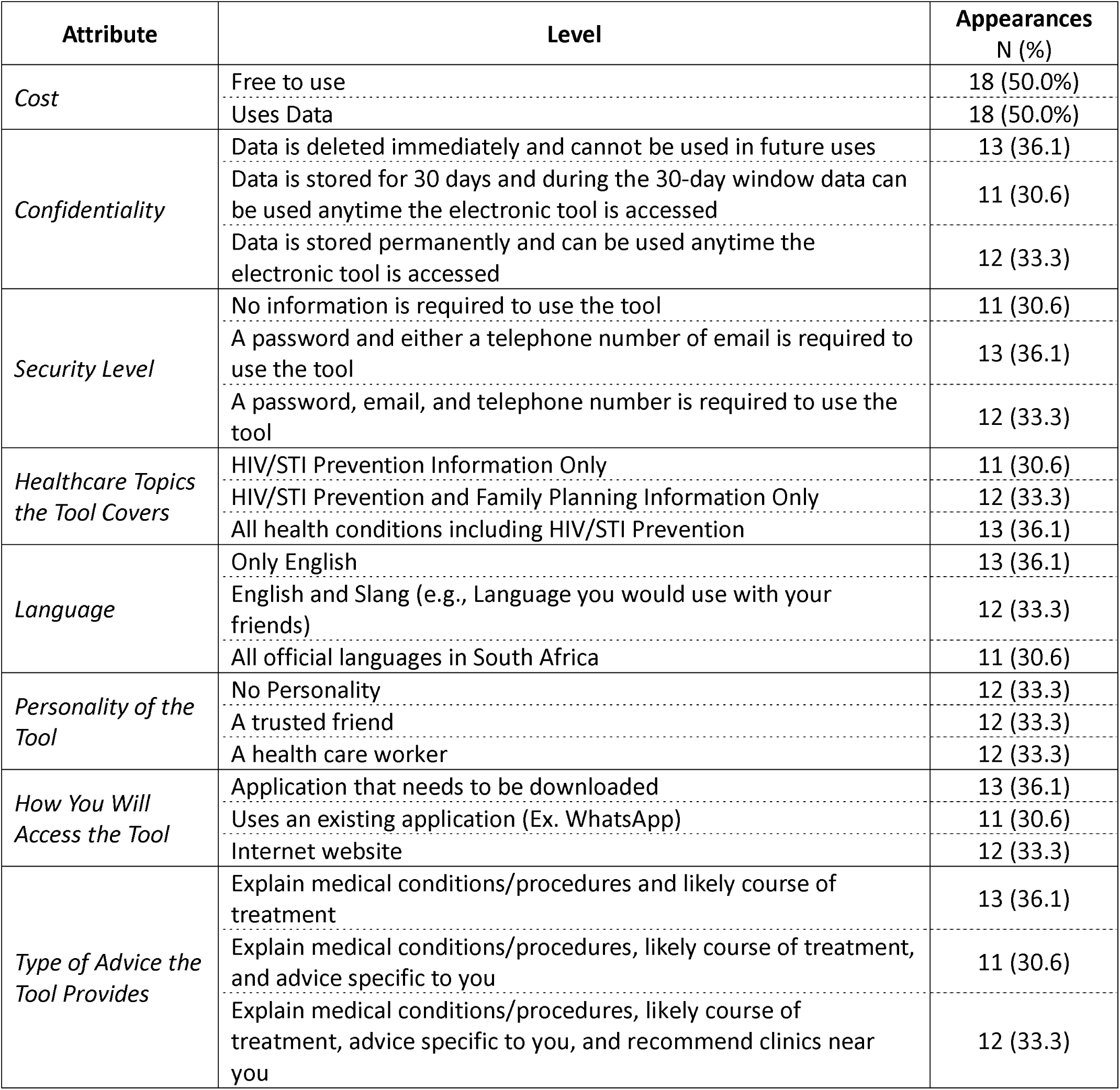
Level Balance

| **Attribute** | **Level** | **Appearances**<br>**N (%)** |
| --- | --- | --- |
| *Cost* | Free to use | 18 (50.0%) |
|  | Uses Data | 18 (50.0%) |
| *Confidentiality* | Data is deleted immediately and cannot be used in future uses | 13 (36.1) |
|  | Data is stored for 30 days and during the 30-day window data can be used anytime the electronic tool is accessed | 11 (30.6) |
|  | Data is stored permanently and can be used anytime the electronic tool is accessed | 12 (33.3) |
| *Security Level* | No information is required to use the tool | 11 (30.6) |
|  | A password and either a telephone number or email is required to use the tool | 13 (36.1) |
|  | A password, email, and telephone number is required to use the tool | 12 (33.3) |
| *Healthcare Topics the Tool Covers* | HIV/STI Prevention Information Only | 11 (30.6) |
|  | HIV/STI Prevention and Family Planning Information Only | 12 (33.3) |
|  | All health conditions including HIV/STI Prevention | 13 (36.1) |
| *Language* | Only English | 13 (36.1) |
|  | English and Slang (e.g., Language you would use with your friends) | 12 (33.3) |
|  | All official languages in South Africa | 11 (30.6) |
| *Personality of the Tool* | No Personality | 12 (33.3) |
|  | A trusted friend | 12 (33.3) |
|  | A health care worker | 12 (33.3) |
| *How You Will Access the Tool* | Application that needs to be downloaded | 13 (36.1) |
|  | Uses an existing application (Ex. WhatsApp) | 11 (30.6) |
|  | Internet website | 12 (33.3) |
| *Type of Advice the Tool Provides* | Explain medical conditions/procedures and likely course of treatment | 13 (36.1) |
|  | Explain medical conditions/procedures, likely course of treatment, and advice specific to you | 11 (30.6) |
|  | Explain medical conditions/procedures, likely course of treatment, advice specific to you, and recommend clinics near you | 12 (33.3) |

**Supplemental Table 3.**
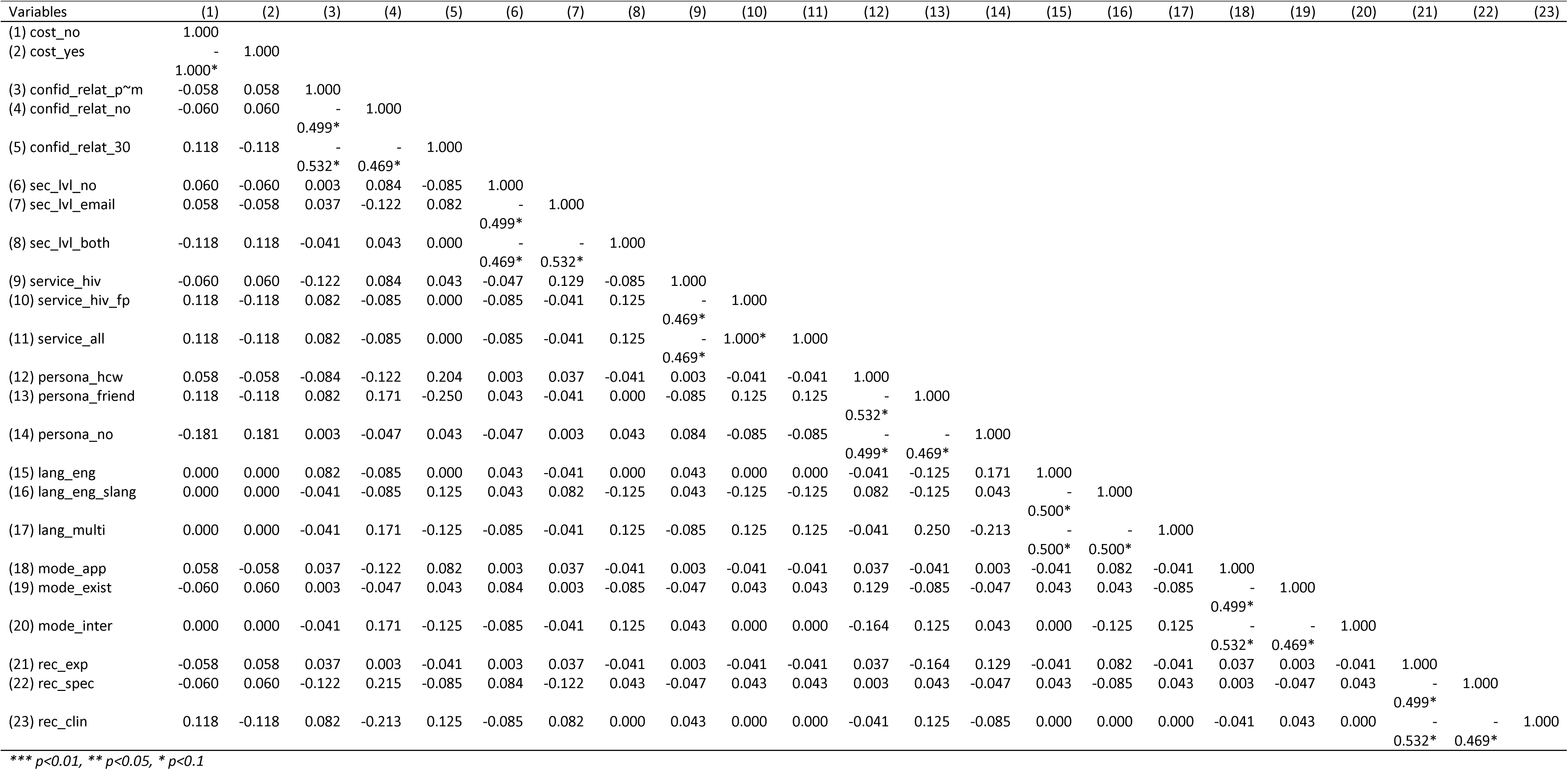
Correlation matrix for orthogonality

**Supplemental Table 4.** Odds Ratios and 95% Confidence Intervals (CI) of Levels in the Primary Analysis

| **Attribute** | **Level** | **Odds Ratio**<br>**(95% CI)** |
| --- | --- | --- |
| *Cost* | Free to use | 1.00 (*Ref*) |
|  | Uses Data | 0.81 (0.74,0.88) |
| *Confidentiality* | Data is deleted immediately and cannot be used in future uses | 1.00 (*Ref*) |
|  | Data is stored for 30 days and during the 30-day window data can be used anytime the electronic tool is accessed | 1.02 (0.90,1.17) |
|  | Data is stored permanently and can be used anytime the electronic tool is accessed | 0.80 (0.70,0.90) |
| *Security Level* | No information is required to use the tool | 1.00 (*Ref*) |
|  | A password and either a telephone number or email is required to use the tool | 1.71 (1.51,1.93) |
|  | A password, email, and telephone number is required to use the tool | 1.71 (1.50,1.94) |
| *Healthcare Topics the Tool Covers* | HIV/STI Prevention Information Only | 1.00 (*Ref*) |
|  | HIV/STI Prevention and Family Planning Information Only | 1.21 (1.06,1.38) |
|  | All health conditions including HIV/STI Prevention | 1.40 (1.24,1.57) |
| *Language* | Only English | 1.00 (*Ref*) |
|  | English and Slang (e.g., Language you would use with your friends) | 1.37 (1.22,1.54) |
|  | All official languages in South Africa | 1.80 (1.60,2.02) |
| *Personality of the Tool* | No Personality | 1.00 (*Ref*) |
|  | A trusted friend | 1.35 (1.18,1.54) |
|  | A health care worker | 1.48 (1.30,1.68) |
| *How You Will Access the Tool* | Application that needs to be downloaded | 1.00 (*Ref*) |
|  | Uses an existing application (Ex. WhatsApp) | 1.49 (1.32,1.68) |
|  | Internet website | 1.14 (1.01, 1.29) |
| *Type of Advice the Tool Provides* | Explain medical conditions/procedures and likely course of treatment | 1.00 (*Ref*) |
|  | Explain medical conditions/procedures, likely course of treatment, and advice specific to you | 1.09 (0.97,1.23) |
|  | Explain medical conditions/procedures, likely course of treatment, advice specific to you, and recommend clinics near you | 1.51 (1.33,1.70) |

**Supplemental Figure 1.**
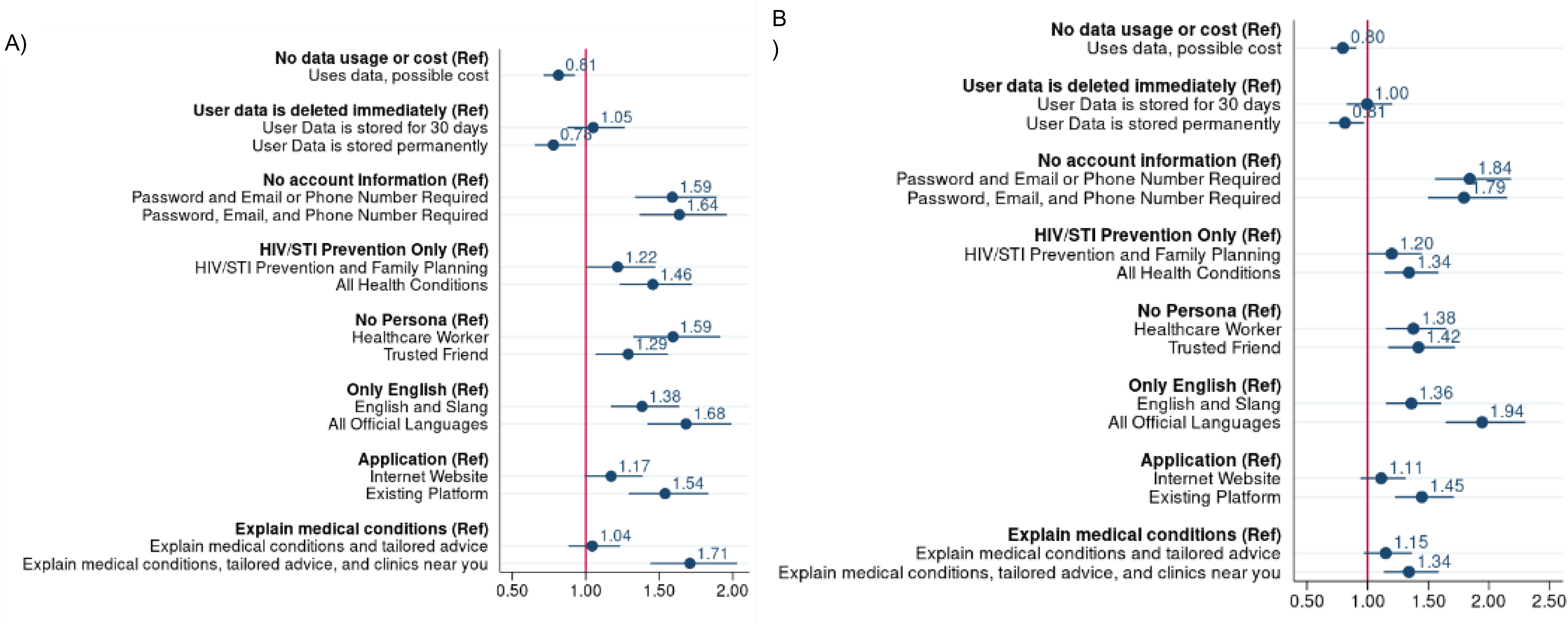
Results from the DCE stratified by A) Females and B) Males

**Supplemental Figure 2.**
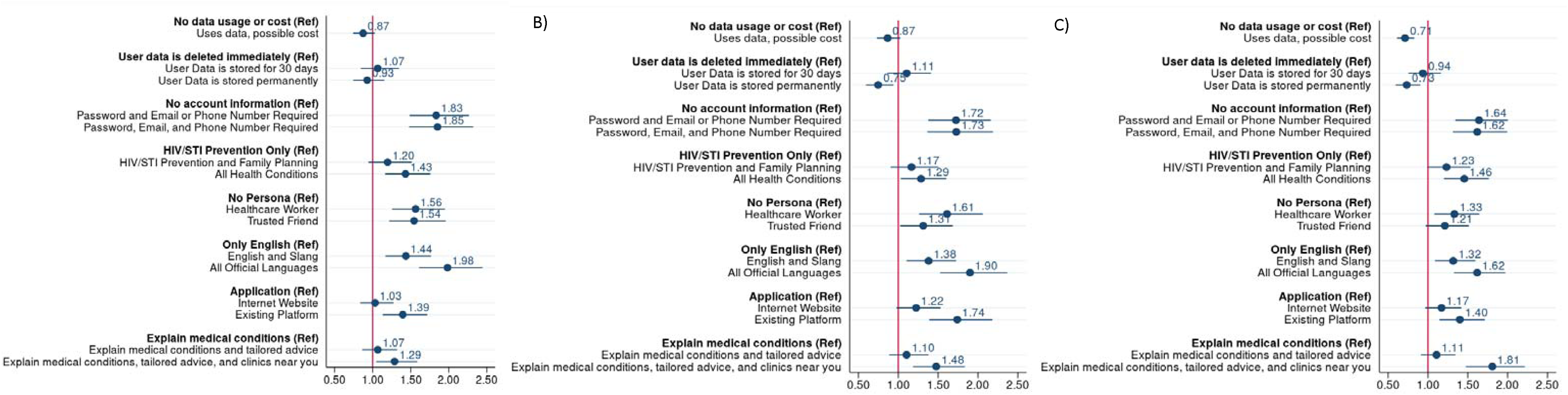
Results from the DCE stratified by A) Low Socioeconomic Status, B) Medium Socioeconomic Status, and C) High Socioeconomic Status

**Supplemental Figure 3.**
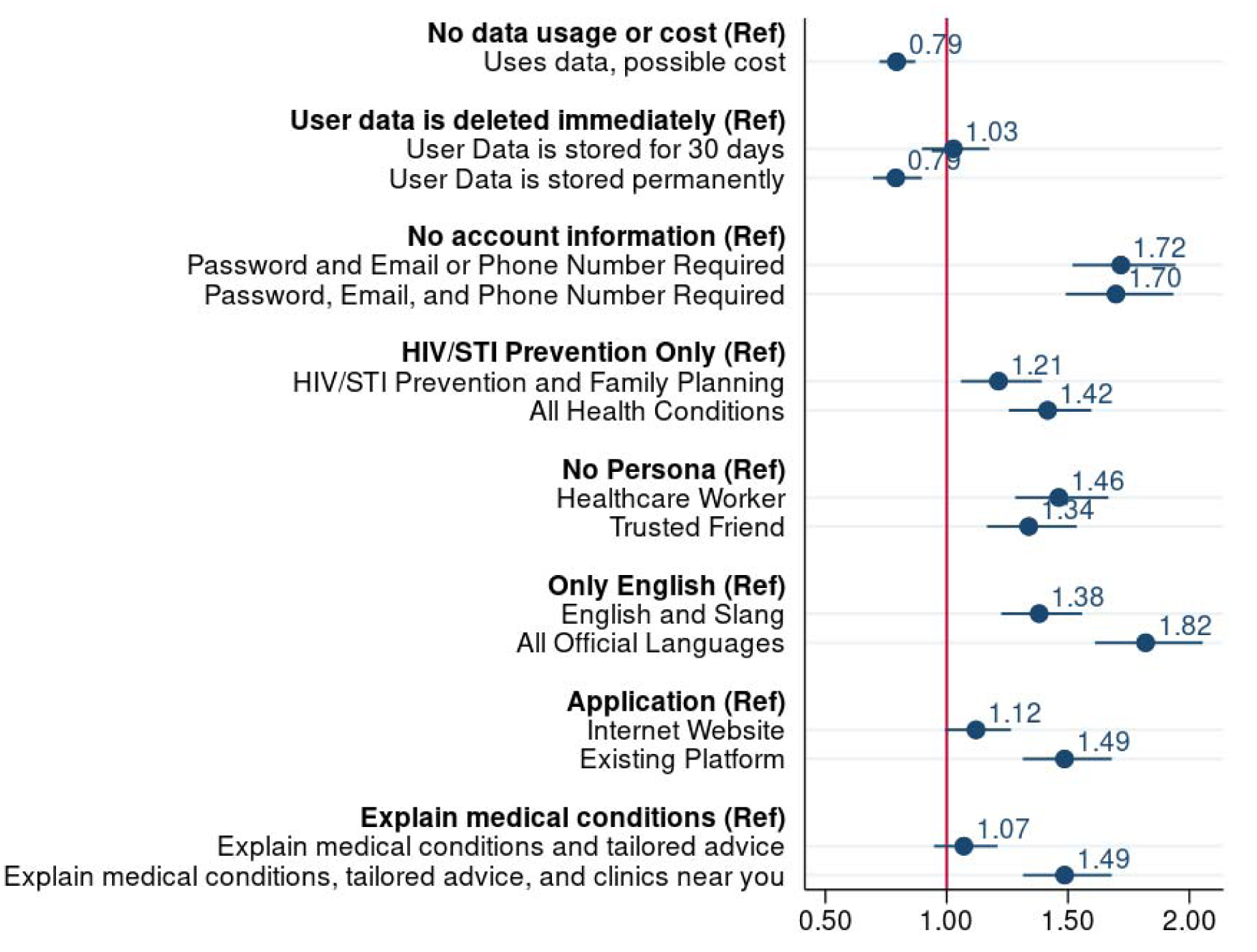
Sensitivity Analysis 1: Excluding participants who completed the DCE in under ten minutes

**Supplemental Figure 4.**
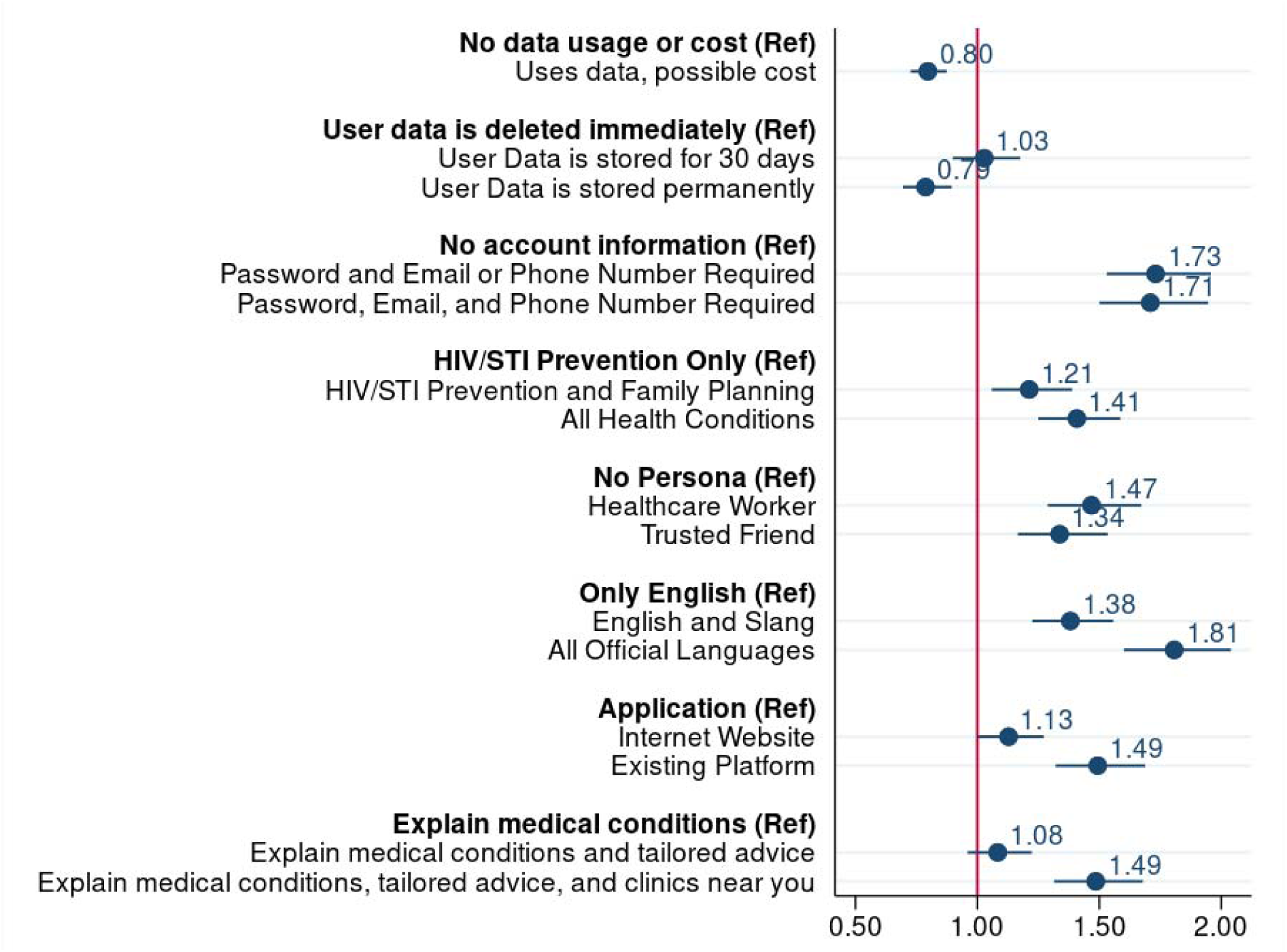
Sensitivity Analysis 2: Excluding the fastest 10% of participants

**Supplemental Figure 5.**
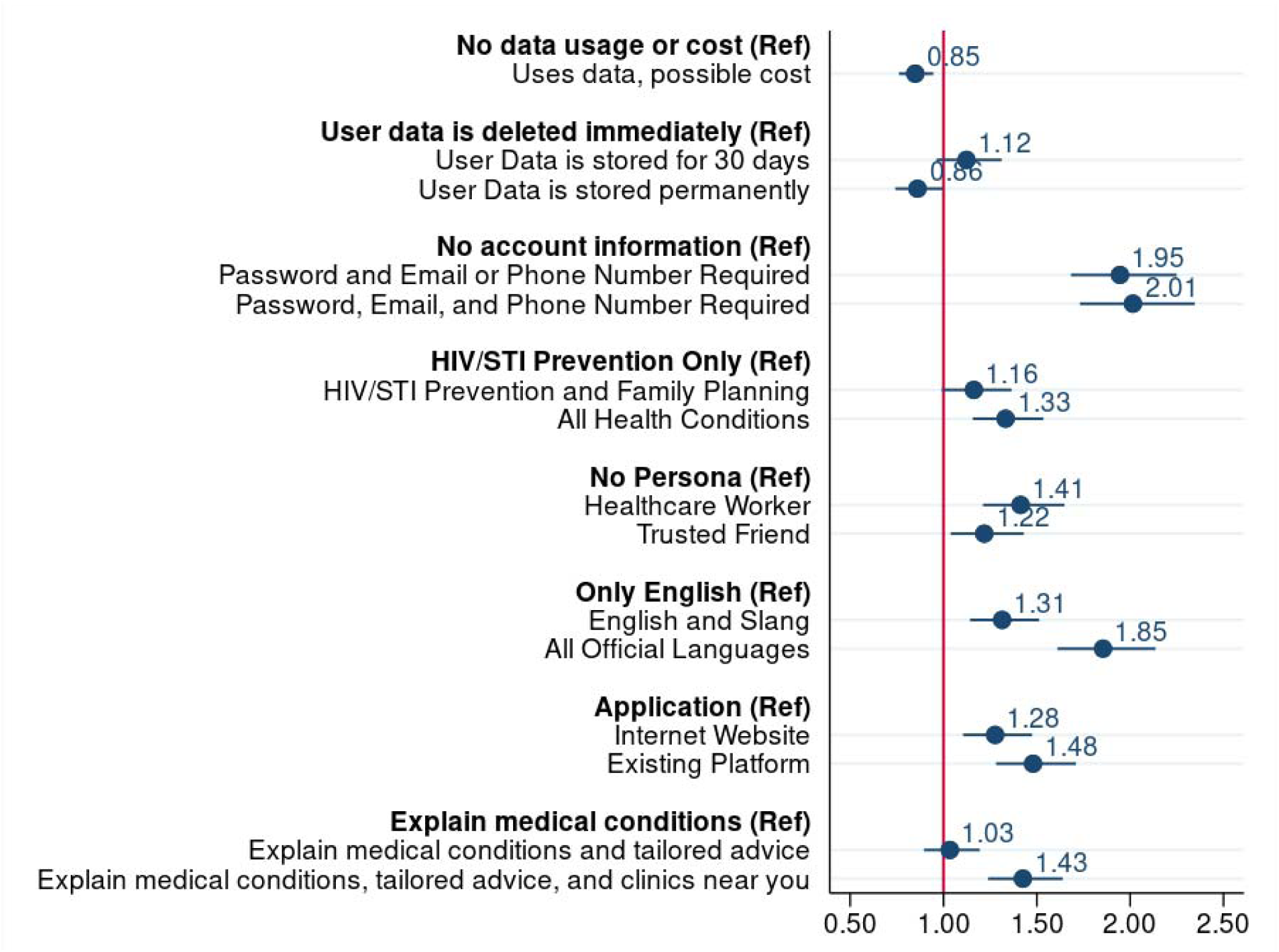
Sensitivity Analysis 3: Excluding participants who did not pass internal consistency

**Supplemental Figure 6.**
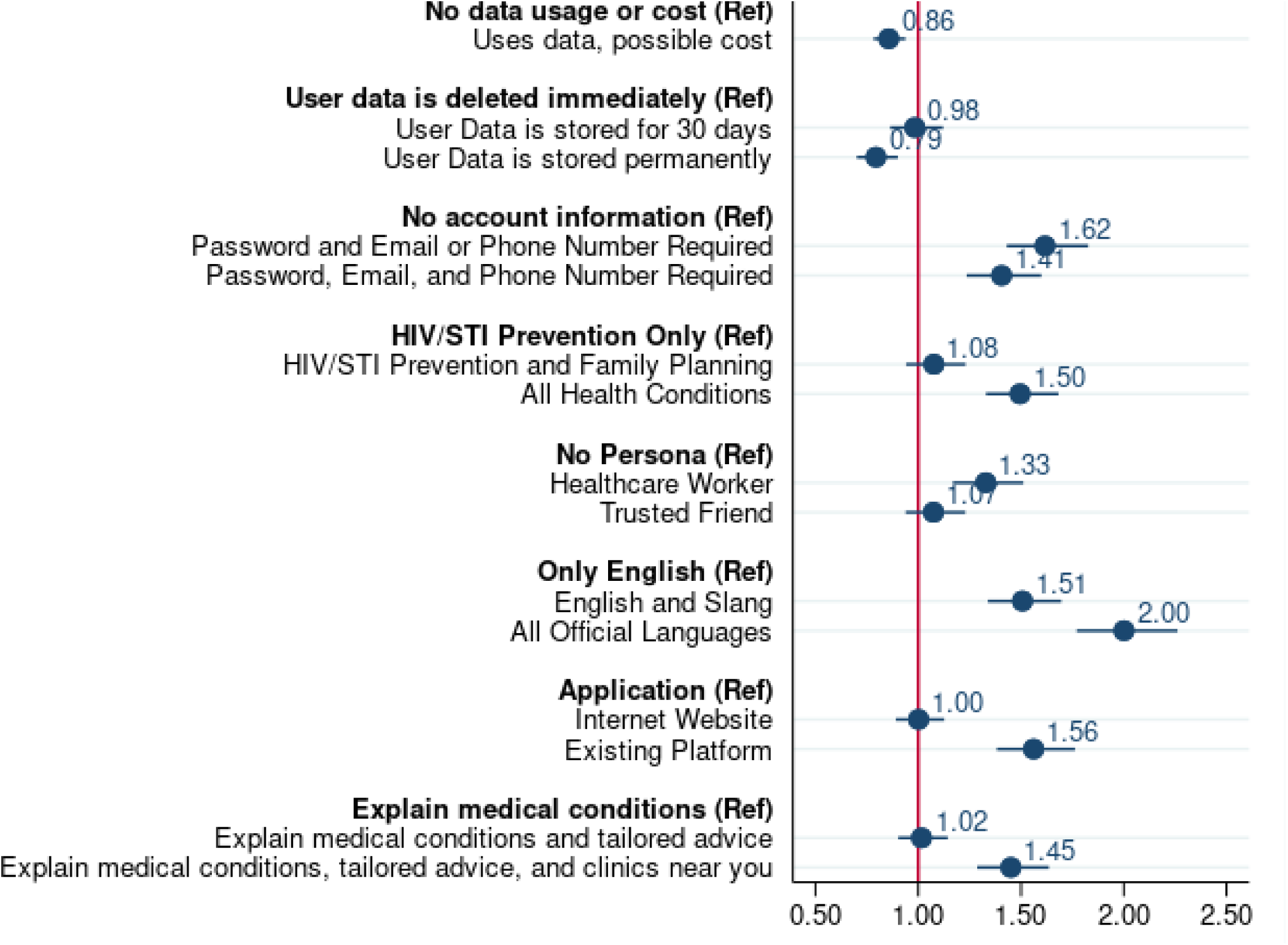
Sensitivity Analysis 4: Using the repeat choice set instead

**Supplemental Text 1.** Overview of the Rank Aggregation Analysis

To create a single “super”-list that is reflective of all thirty responses, we conducted a rank aggregation using the Cross-Entropy Monte Carlo algorithm. At a high-level the algorithm uses a series of matrices to determine what the ideal list is. This involves four steps: 1) within each matrix, each cell has a uniform multinomial probability, 2) at each stage, a random sample of the matrices are generated via truncated multinomial sampling based on the cell probabilities of the select matrices. Third, based on the current sample for that stage and the list that is subsequently generated, the multinomial cell probabilities generated in Step 1 are updated with the goal of reducing the variability for the next random sample. Step 3 is repeated until convergence (Step 4) is achieved in which the list that is generated does not change for a given number of samples. The smaller the number of iterations and optimal value it takes to achieve convergence the better. It is important to note that if one were to run the algorithm several times, the “super”-list generated may vary slightly depending on the seed set and if the sample size is small.

